# Magnitude of Psychological Distress among Medical and Non-medical Students during the Late Phase of COVID-19 Pandemic in West Bengal: A Cross-sectional Study

**DOI:** 10.1101/2023.07.30.23293045

**Authors:** Udisa Das, Arunima Ganguly, Dibakar Haldar, Asish Mukhopadhyay

## Abstract

**Background:** COVID-19 led to drastic changes worldwide which has affected mental health especially, of the vulnerable student population.

**Aim:** This study aimed to assess psychological distress due to COVID-19 in students during the late phase of pandemic and to establish correlation of academic course, socio-demographics and knowledge-attitude-practices (KAP) with depression and anxiety. An online cross-sectional survey was conducted among medical and non-medical students in Kolkata, from March to April 2022.

**Methods:** A cross-sectional survey was done using purposive and snowball sampling methods. Survey questionnaire was circulated via Google forms through social media. It included Patient Health Questionnaire-9, Generalized Anxiety Disorder-7, Fear of COVID-19 scale 2020, KAP regarding COVID-19 and socio-demographics. Data were analyzed using appropriate statistical methods with the Statistical Package for the Social Sciences (Version 22.0). P value of less than 0.05 was considered significant.

**Results:** Total of 442 responses were received. After excluding incomplete responses, the final sample comprised 219 medical and 202 non-medical students. Majority of the participants were male (58.67%). Overall prevalence of depression was 58.42% in non-MBBS and 81.73% in MBBS students. Whereas, the overall prevalence of anxiety was 50.99% in non-MBBS and 76.25% in MBBS students. MBBS participants had significantly better scores for knowledge and attitude (p=0.000 in both). Depression was higher in those with previous history of psychiatric illness (p=0.015). Anxiety was influenced by residence (p=0.003), mode of travel to college (p=0.002), history of relatives or friends affected by COVID-19 (p= 0.005).

**Conclusion:** Prevalence of depression and anxiety in college students, especially among medical students, was higher in present study mainly due to long-term indirect effects of the pandemic. This calls for employing student wellness activities and provision of better mental health services across colleges in India.

**Key Messages:** Depression and anxiety was observed to be higher especially among medical students. Previous history of psychiatric illness was found to be a correlate of depression. Anxiety was revealed to be influenced by residence, mode of travel to college, and history of family, friends or relatives affected by COVID-19.

## INTRODUCTION

COVID-19 began as an epidemic in China in December 2019 and was declared a pandemic by World Health Organisation in March 11, 2020.^1^ Due to severe transmissibility of the SARS-COV2 virus and its adverse outcomes in some cases, the main focus has been on physical effects of pandemic. This led to drastic lockdowns and strict quarantine measures worldwide. India implemented its first lockdown in four phases from 25th March, 2020 to 31st May, 2020.^2^ The second wave of pandemic began in March, 2021. Since its discovery, fear of the virus has greatly affected mental health of individuals.^3^ Studies conducted amongst general population identified huge rise in symptoms of depression and anxiety.^4^ Meanwhile, medical students are already more prone to develop psychological distress because of highly demanding medical curriculum and other factors.^5^ By March 2022, due to widely successful vaccination drive in India, 70% of the population had received at least one dose of vaccine. Thus, gradual opening up protocol was followed.^6^ Schools, colleges and offices returned to offline mode. Public transports, restaurants and tourist attractions opened up. Given the current situation being greatly different from that during lockdown, factors determining mental health of the student population are also expected to change.

There are few studies conducted in India regarding impact of COVID-19 on mental health. Most of them focus on the initial phase of pandemic during early lockdown period and show different results for prevalence of depression and anxiety. In a study conducted in Karnataka,^7^ depression in girls was found to be more than that in boys and anxiety did not differ between genders whereas, in a study conducted in Chennai,^8^ depression increased in males and anxiety increased in females due to COVID-19. Another study conducted in West Bengal showed a significantly different prevalence of anxiety.^9^ Also, in West Bengal very few studies have been conducted to assess mental health of university students during the pandemic. Probably, studies comparing mental distress among medical and non-medical students in West Bengal in the late phase of the pandemic (when gradual opening-up protocols were being followed after the second wave of the pandemic) have not been conducted. The objectives of this study were:

1. To estimate the prevalence of Depression and Generalised Anxiety Disorder (GAD) among medical and non-medical students during the late phase of COVID-19 pandemic
2. To assess the Knowledge, Attitude and Practice (KAP) regarding COVID-19 between medical and non-medical students
3. To determine the relation between Depression, GAD and KAP regarding COVID-19 among the participants
4. To find out the correlates of psychological distress among the participants

This study focused on the medical and non-medical students, who were already under tremendous stress due to disruption of education and now due to the possible chance of getting more exposure caused by resumption of offline classes. This would help assess the long-termpsychological burden of COVID-19 pandemic after two years since the first case was detected, if any and also help the college authorities to develop strategies to improve the mental wellbeing and thereby, learning ability of students.

## METHODS

This study was a cross-sectional survey conducted among medical and non-medical students (general and engineering) of Kolkata, during late phase of the pandemic. Study participants included third year undergraduate medical students (having clinical ward classes) and third year undergraduate non-medical students. Purposive sampling was employed to choose the study participants with the above-mentioned specific characteristics for fair comparison. Snowball sampling was used to enable greater reach. Using the prevalence (35.5%) explored by relevant research as ‘P’,^8^ sample size has been calculated using formula n= [Z^2^PQ]/L^2^, where Z=1.96 at 95% confidence interval (CI), Q=complement of P =100-P and L=7, absolute error around the reported prevalence. The sample size for the study has been estimated to be (3.84*35.5*64.5)/7^2^=180. Being an online survey, after adding 20% non-response rate the sample size was 216. So, 220 medical students along with 220 non-medical students were considered adequate for the study.

Participants were asked to fill online Google forms distributed by social media platforms like WhatsApp and Facebook. Data collection was conducted from 8th March, 2022 till 6th April, 2022. Forms were accepted till responses reached 220 for both study groups. Total of 442 responses were received. After excluding incomplete and inappropriate responses, the final sample comprised 421 participants of whom 219 were medical and 202 were non-medical students.

A validated and pretested questionnaire containing information pertaining to socio-demographics was used.

To assess depressive symptoms, participants completed the nine-item Patient Health Questionnaire (PHQ-9).^10^ PHQ-9 is a self-reported scale used to diagnose major and subthreshold depression. Participants indicated how frequently they experienced depressive symptoms over the past two weeks on a four-point Likert scale, from 0 “not at all” to 3 “nearly every day”. The total score range is 0–27 which determines the severity of depression. It is interpreted as normal (0–4), mild (5–9), moderate (10–14), moderately severe (15-19) and severe (20-27) depression.

The seven-item Generalized Anxiety Disorder (GAD-7) was used to assess anxiety symptoms.^11^ Participants indicated how frequently they experienced symptoms of anxiety over the last two weeks on a four-point Likert scale from 0 “not at all” to 3 “nearly every day”. Total score of the participants ranges from 0-21. The severity of symptoms of anxiety is interpreted as normal (0–4), mild (5–9), moderate (10–14), and severe (15–21) anxiety.

To assess Fear of COVID-19, Fear of COVID-19 Scale, 2020 (FCV-19S) was used.^12^ This is a reliable, valid self-report scale developed recently to assess fear of COVID-19 pandemic. A study conducted by Ahorsu et.al^12^ found reliability values like, internal consistency (alpha = 0.82) and test-retest reliability (ICC = 0.72) for this scale which were acceptable. The participants indicated their level of agreement with the statements using a seven-item questionnaire on a five-point Likert scale,from 1 to 5Answers included “strongly disagree,” “disagree,” “neutral” “agree” and “strongly agree”. Total score was calculated by adding up each item score (ranging from 7 to 35). Higher score (score >18) corresponded to greater fear.^15^

The pre-validated Knowledge, Attitude and Practices regarding COVID-19 questionnaire adopted from relevant study,^13^ was modified by subject matter experts, to suit current study sample. The Knowledge section consisted of six questions related to mode of transmission, symptoms, management options and preventive strategies. The questions had answers as “Yes”, “No” and “Do not know”. Participants who answered 50%, two-third or more and less than 50% of the questions correctly, were respectively graded as “Average”, “Good” and “Poor”. The Attitude section had four questions related to possibility, severity of infection, attitude towards practicing personal hygiene and avoiding crowded places. These were graded by five-point Likert scale. Those who attained median Attitude score of 11, more than 11 and less than 11 were respectively, graded as “Average”, “Good” and “Poor”. The Practice section had three questions related to exercising preventive strategies which were graded by four-point Likert scale. Participants who attained median Practice score of ten, more than ten and less than ten were graded as “Average”, “Good” and “Poor”, respectively.

Collected data was compiled in Microsoft Excel. The data was summarised and analysed using appropriate statistical methods. Continuous data was described by mean, median, standard deviation (SD), interquartile range (IQR) etc. and categorical data by proportion and percentage. Normality of data set was checked by charts like histogram, stem-leaf, P-P and Q-Q plot and Shapiro-Wilk normality test. Inferential statistical tests like ‘Unpaired t’ test (for normally distributed data), Pearson correlation coefficient (r)/ Mann-Whitney U test (for skewed data) was used for continuous variables. The Statistical Package for Social Science (SPSS Version 22.0) was used for analysis. P-value of less than 0.05 was considered as significant.

### Ethical Approval

The study was carried out after obtaining approval of the Institutional Ethics Committee of Nilratan Sircar Medical College and Hospital, Kolkata on 23rd February, 2022 with Memo no: NRSMC/IEC/03/2022 and conducted according to the World Medical Association Declaration of Helsinki on Ethical Principles for Medical Research Involving Humans. Informed online consent was obtained from each study participant after explanation of the study and confirming confidentiality.

## RESULTS

Total of 442 responses were received. After excluding incomplete and inappropriate responses, the final sample comprised 421 participants of whom 219 were medical and 202 were non-medical students. Majority of the participants were male (58.67%). Mean age was 22.42±0.99 (mean±SD) years.Other socio-demographic characteristics are depicted in **Table 1**.

**Table 1.** Distribution of Participants According to Socio-demographic Characteristics, West Bengal, 2022.

| Variable | Attribute | Number | % |
| --- | --- | --- | --- |
| Gender | Male | 247 | 58.67 |
|  | Female | 174 | 41.33 |
| Age in years (Mean±SD) |  | 22.42±0.99 |  |
| Course | MBBS | 219 | 52.02 |
|  | Non-MBBS | 202 | 47.98 |
| Residence during Late Phase of Pandemic | Home | 203 | 48.22 |
|  | Hostel | 162 | 38.48 |
|  | Paying Guest | 56 | 13.3 |
| Mode of Travel to College | Hostel-boarder | 162 | 38.48 |
|  | Public Transport | 204 | 48.46 |
|  | Hired Car | 27 | 6.41 |
|  | Own Car | 28 | 6.65 |
| History of Addiction | Alcohol | 6 | 1.43 |
|  | Tobacco | 28 | 6.65 |
| Previous History of Psychiatric Illness | Depression | 5 | 1.19 |
|  | Anxiety neurosis | 3 | 0.71 |
|  | Obsessive Compulsive Disorder | 4 | 0.95 |
| Presence of comorbidities | Yes | 81 | 19.23 |
|  | No | 340 | 80.76 |
| If any family member/ close relative or friend, suffered/died from COVID-19 | Yes | 250 | 59.38 |
|  | No | 171 | 40.62 |
| If any family member is working in health care system | Yes | 93 | 22.09 |
|  | No | 328 | 77.91 |

Overall prevalence of depression (PHQ-9 score>4) was 58.42% in non-MBBS and 81.73% in MBBS students. Whereas, the overall prevalence of anxiety (GAD-7 score >4) was 50.99% in non-MBBS and 76.25% in MBBS students. The PHQ-9 score (mean+/-SD) was 7.62 +/- 5.86 for the non-MBBS participants and 8.37 +/- 5.16 for the MBBS participants, respectively with p=0.017 (Mann-Whitney U: 19136.000).**(Figure 1)**

**Figure 1.**
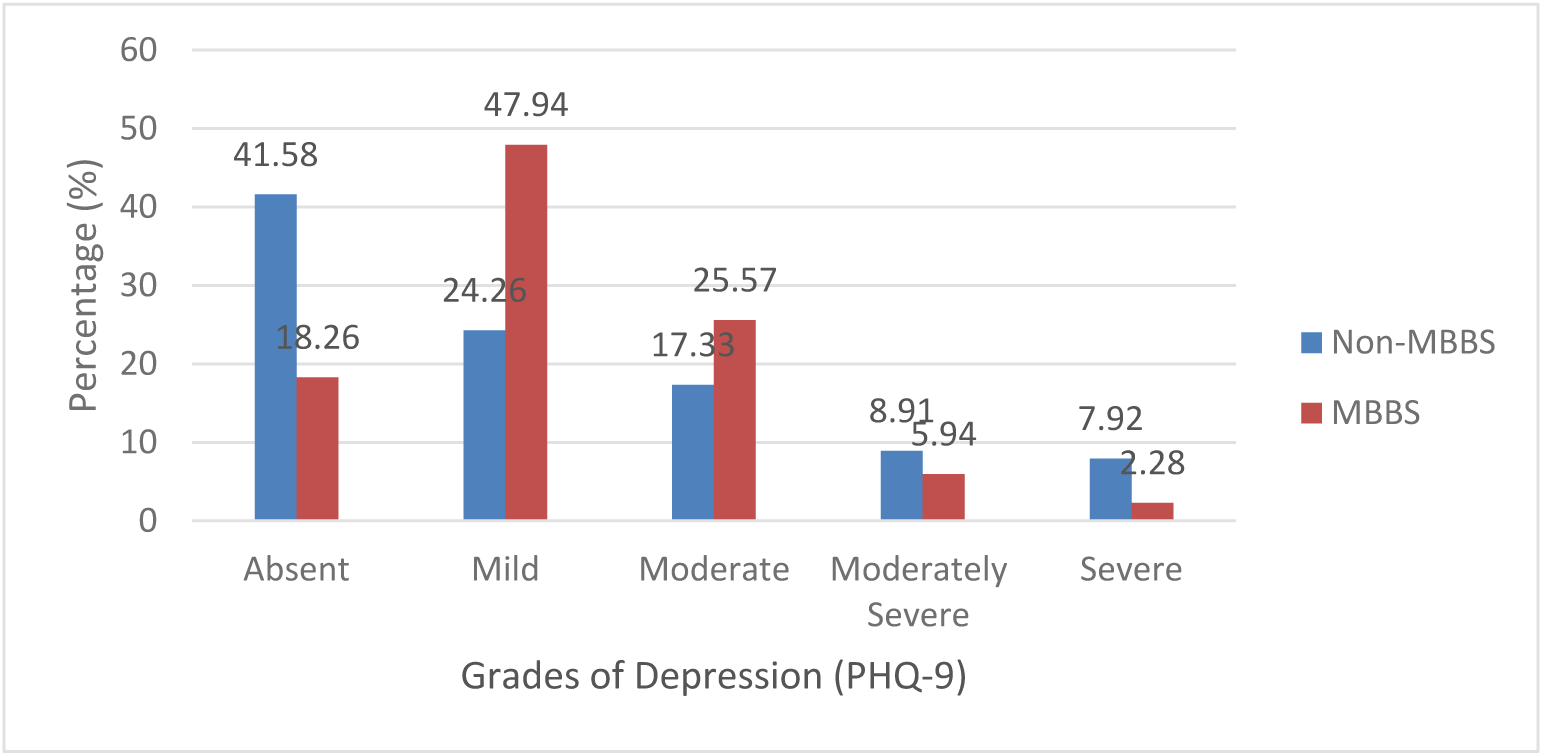
Distribution of Participants According to Academic Course and Grades of Depression(PHQ-9), West Bengal, 2022

The GAD-7 score (mean+/-SD) was 7.20 +/- 5.63 for the non-MBBS participants and 7.96 +/- 4.65 for the MBBS participants, respectively with p=0.008 (Mann-Whitney U: 18801.000).**(Figure 2)** According to FCV-19S, non-MBBS and MBBS participants had scores of 14.93±7.3 (mean±SD), 12.00(8.00) [median (IQR)] and 13.63 ± 5.07 (mean±SD), 13.00 (5.00) [median (IQR)]; respectively. MBBS participants were found to have higher score of knowledge regarding COVID-19 than their non-MBBS counterparts which were, 4.24±1.04 (mean±SD)and 3.80±1.28 (mean±SD), respectively (Mann-Whitney U & p-values 16396.00, 0.000). Attitude scores were 10.64±2.26 (mean±SD) vs. 11.63±1.93 (mean±SD) for the non-MBBS vs. MBBS participants, respectively which was statistically significant (Mann-Whitney U & p-values 15207.500, 0.000). Practice scores for non-MBBS vs. MBBS participants were 9.85±1.92 (mean±SD) vs. 9.44±1.74 (mean±SD), respectively, which had borderline statistical significance (Mann-Whitney U & p-values 19714.00, 0.049).**(Table 2)**

**Figure 2.**
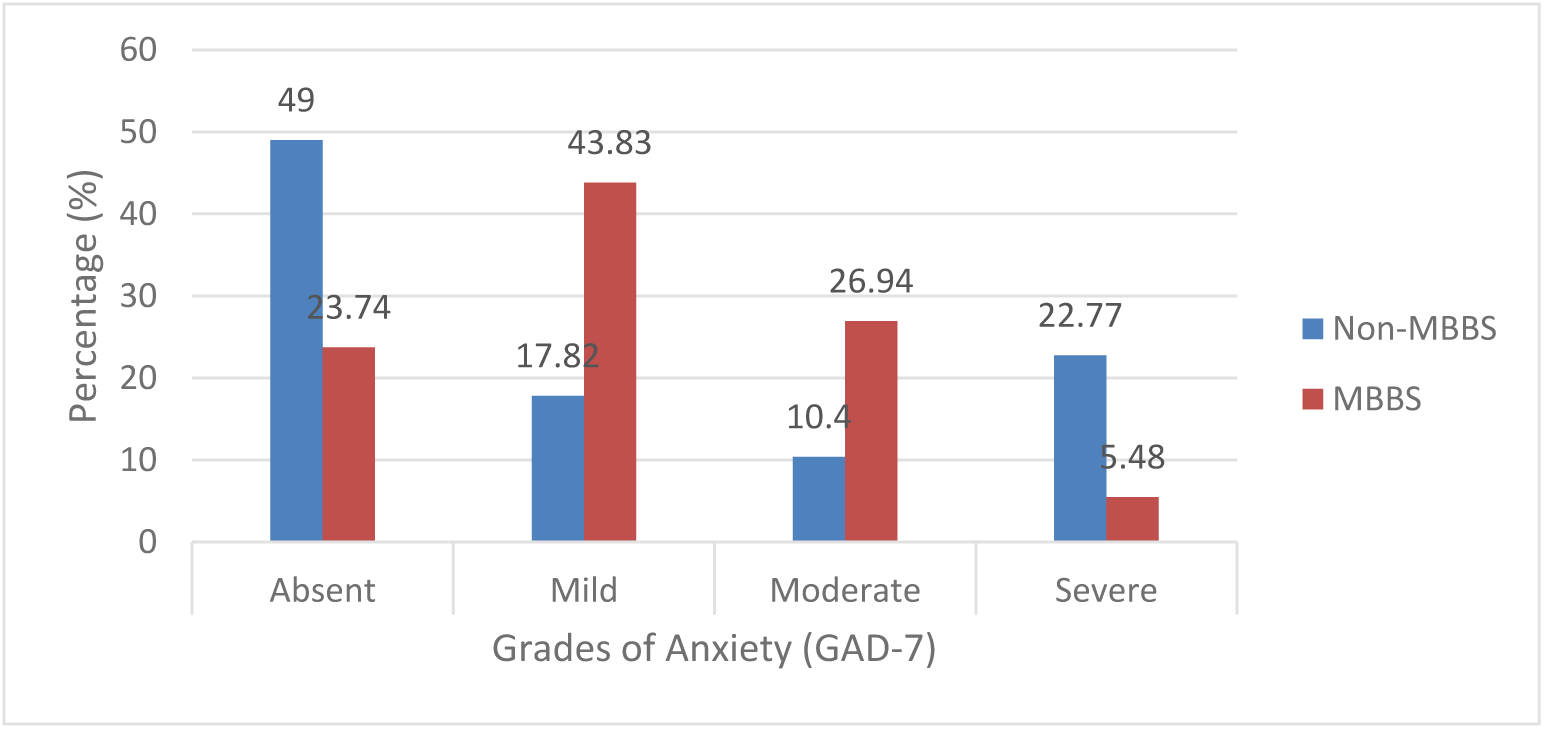
Distribution of Participants According to Academic Course and Grades of Anxiety(GAD-7), West Bengal, 2022

**Table 2.** Distribution of Participants according to Academic Course and Knowledge, Attitude and Practices regarding COVID-19, West Bengal, 2022.

|  | Course | Score<br>(mean±SD) | Median (IQR) | Mann-Whitney<br>U | P- value |
| --- | --- | --- | --- | --- | --- |
| Knowledge | Non-MBBS | 3.80±1.28 | 4.0 (1.0) | 16396.00 | **0.000** |
|  | MBBS | 4.24±1.04 | 5.0 (2.0) |  |  |
| Attitude | Non-MBBS | 10.64±2.26 | 10.0 (3.0) | 15207.50 | **0.000** |
|  | MBBS | 11.63±1.93 | 12.0 (3.0) |  |  |
| Practice | Non-MBBS | 9.85±1.92 | 9.50 (3.0) | 19714.00 | 0.049 |
|  | MBBS | 9.44±1.74 | 10.0 (3.0) |  |  |

Significant results have been marked in bold
IQR= Interquartile range

Grades of Attitude were found to significantly influence Depression scores (p=0.011). Participants having average attitude towards COVID-19 had a greater mean score of depression than those having poor attitude and also those having good attitude, which was found to be significant in Post-Hoc analysis with p-values 0.009 and 0.036, respectively. Attitude grades were also found to significantly influence GAD-7 scores with a p-value of 0.045.**(Table 3)**

**Table 3.** Distribution of Participants as per Knowledge, Attitude and Practices along with their Depression, GAD and Fear of COVID-19 Scores, West Bengal, 2022.

| Grades | PHQ-9<br>Score<br>(mean±SD) | H<br>(Kruskal<br>Wallis<br>ANOVA),<br>p value | GAD-7<br>Score<br>(mean±SD) | H<br>(Kruskal<br>Wallis<br>ANOVA),<br>p value | Fear of<br>COVID-19<br>Score<br>(mean±SD) | H<br>(Kruskal<br>Wallis<br>ANOVA),<br>p value |
| --- | --- | --- | --- | --- | --- | --- |
| Knowledge |  |  |  |  |  |  |
| Poor | 6.98±0.85 | 2.753,<br>0.253* | 7.57±0.77 | 0.159,<br>0.923* | 13.66±0.96 | 0.780,<br>0.677* |
| Average | 8.08±0.74 |  | 7.53±0.71 |  | 14.89±0.89 |  |
| Good | 8.14±0.31 |  | 7.62±0.29 |  | 14.21±0.34 |  |
| Attitude |  |  |  |  |  |  |
| Poor | 7.56±0.39 | 8.984,<br>**0.011\*** | 6.98±0.36 | 6.185,<br>**0.045\*** | 14.02±0.42 | 0.227,<br>0.893* |
| Average | 9.75±0.71 |  | 8.27±0.57 |  | 13.62±0.75 |  |
| Good | 7.94±0.43 |  | 8.03±0.42 |  | 14.69±0.52 |  |
| Practice |  |  |  |  |  |  |
| Poor | 7.76±0.37 | 2.797, | 7.60±0.36 | 2.804, | 13.97±0.42 | 2.909, |
| Average | 8.81±0.61 | 0.247* | 8.02±0.50 | 0.246* | 13.20±0.57 | 0.234* |
| Good | 7.89±0.50 |  | 7.34±0.48 |  | 15.31±0.62 |  |

\*df=2
Significant results have been marked in bold

Depression was found to be significantly higher in those having previous history of psychiatric illness (p=0.015). Anxiety differed significantly according to residence (p=0.003), mode of travel to college (p=0.002), history of family member/close relative/friend suffering or dying from COVID-19 (p=0.005). Those travelling by own car had greater mean score of anxiety than those travelling by public transport as per Post-Hoc analysis. No significant relationship was found between fear of COVID-19 and above factors.**(Table 4)**

**Table 4.** Distribution of Participants according to Depression, Generalised Anxiety Disorder, Fear of COVID-19 and Socio-demographics, West Bengal, 2022.

| Variable | Attributes | PHQ-9 Score<br>(mean±SD) | p value | GAD-7<br>Score<br>(mean±SD) | p value | Fear of<br>COVID-19 Score<br>(mean±S<br>D) | p<br>value |
| --- | --- | --- | --- | --- | --- | --- | --- |
| Gender | Male | 7.72(5.42) | 0.135* | 7.26(5.08) | 0.065* | 14.10(6.2<br>3) | 0.524* |
|  | Female | 8.38(5.64) |  | 8.03(5.19) |  | 14.46(6.3<br>5) |  |
| Place of<br>Residence | Home | 7.98(5.67) | 0.116<sup>†</sup> | 7.53(5.29) | **0.003<sup>†</sup>** | 14.19(6.5<br>9) | 0.546<sup>†</sup> |
|  | Hostel | 8.41(5.38) |  | 8.34(5.09) |  | 14.42(6.0<br>4) |  |
|  | Paying<br>Guest | 7.00(5.37) |  | 5.79(4.37) |  | 13.93(5.8<br>3) |  |
| Mode of<br>Travel to | Hostel-boarder | 8.30(5.31) | 0.329<sup>‡</sup> | 8.33(5.05) | **0.002<sup>‡</sup>** | 14.48(6.0<br>9) | 0.108<sup>‡</sup> |
| College | Public Transport | 7.58(5.56) |  | 6.81(5.11) |  | 13.83(6.24) |  |
|  | Hired Car | 8.56(4.88) |  | 6.85(4.39) |  | 13.52(5.44) |  |
|  | Own Car | 8.96(6.84) |  | 9.82(5.65) |  | 16.79(7.72) |  |
| Previous History of Psychiatric Illness | No | 7.90(5.50) | **0.015\*** | 7.57(5.14) | 0.522* | 14.31(6.29) | 0.331* |
|  | Yes | 11.07(5.28) |  | 8.47(5.55) |  | 12.87(5.58) |  |
| If any family member/ close relative or friend, suffered/died from COVID-19 | Not suffered/died | 7.59(5.42) | 0.134* | 6.75(4.92) | **0.005\*** | 14.02(5.92) | 0.771* |
|  | Suffered/died | 8.30(5.57) |  | 8.18(5.23) |  | 14.42(6.51) |  |
| If any family member is working in health care system | No | 7.76(5.37) | 0.073* | 7.45(5.15) | 0.193* | 14.11(6.26) | 0.207* |
|  | Yes | 8.89 (5.95) |  | 8.12(5.15) |  | 14.76(6.31) |  |

\*According to Mann-Whitney U test, †According to Kruskal Wallis ANOVA at df=2, ‡According to Kruskal Wallis ANOVA at df=3, Significant results have been marked in bold

## DISCUSSION

In this study, prevalence of mild to severe symptoms of depression and anxiety in MBBS students was observed to be higher as compared to another study.^14^ This variation might be due to methodological and socio-cultural differences. Comparable values of prevalence of depression but a higher prevalence of anxiety among non-MBBS students was found in a study conducted among university students in India.^15^

The mean PHQ-9 and GAD-7 scores for non-MBBS participants were significantly lower than that of MBBS participants. This might be due to offline mode of classes, wards and examinations which had already started for MBBS students during the study period. Also, higher prevalence of psychological distress might be because, medical students are subjected to various challenges like, demanding curriculum, long study hours, fear of failure, as reported in another study.^5^

The 2016 National Mental Health Survey reported 2.7% prevalence of depressive disorder and 3.1% prevalence of anxiety in Indian population.^16^ Prevalence of depression and anxiety were found to be 27.2% and 33.8% respectively, among medical students before onset of COVID-19.^17,18^

A study conducted during lockdown in India among non-medical students found 85.51% and 62% of students had symptoms of depression and anxiety.^19^ Similar findings were reported among university students in Bangladesh.^20^

In our study, percentage of depression and anxiety were found to be lower than that during lockdown for non-MBBS students. , Among medical students, depression was found to be lower than that during lockdown.^19^ This decline in distress is supported by other studies conducted after first lockdown in India.^14,15^ The progressive decline in prevalence of distress is in accordance with another longitudinal study.^21^ However, levels of anxiety and depression were still higher than that before the pandemic among both MBBS and non-MBBS students, which is compliant with another study.^3^

MBBS students had higher knowledge scores (p=0.000) than their non-MBBS counterparts. This maybe because COVID-19 has lately been incorporated in the MBBS curriculum in India.

Non-MBBS students had a lesser mean score of attitude than their MBBS counterparts (p=0.000). The knowledge of the non-MBBS participants was based on mass-media which, at the time of this study, showed decreased number of COVID-19 cases. Moreover, they had no clinical ward exposure. Thus, less perceived possibility and severity of infection might have led to lesser mean scores of attitude.

Although, mean score of practice was slightly higher among the non-MBBS participants, it showed almost no statistical significance (p=0.049). This is because, practices like wearing masks and hand-washing had been integrated into the daily lives of the population for two years since start of the pandemic. Social demand and legal enforcement might be the likely explanations of similar practice scores among both study groups.

At the start of the pandemic, little was known about transmission, pathogenesis, complications of COVID-19 and there was high amount of unverified information, leading to uncertainty that may have led to strict protective measures and thus, higher attitudes and practices despite poor knowledge, as found in a study conducted in Indonesia during early stages of the pandemic.^22^ During this study conducted in late phase of the pandemic, extensive research followed by mass media campaigning has led to increased knowledge scores. The difference in attitude and practice with earlier studies might be due to decrease in hospitalisation and deaths due to COVID-19 and the pan-India vaccination drive, reducing associated fear. Two years have passed since the start of the pandemic and people have become complacent with their practices, leading to relatively lower practice scores, supported by another study.^23^

Participants having average attitude towards COVID-19 had greater mean score of depression than those having good attitude. This may be because most of the participants having good attitude also showed average to good practices, thereby having less perceived risk of contracting COVID-19, leading to lower depression scores. Positive attitude towards COVID-19 has been found to negatively correlate with psychological distress.^24^ However, poor attitude during the late phase of pandemic also had significantly less mean scores of depression. This is most probably because those participants were reckless regarding the pandemic and so experienced a false sense of wellbeing and denial thereby, less symptoms of depression.

Depression was found to be higher in those having prior history of psychiatric illness, compliant with a study in Pakistan.^3^

Participants staying in hostel had significantly greater mean scores of anxiety. This might be because proper attention is often not paid to maintenance of hygiene and sanitation in hostels. Moreover, it is not expected that all hostel-boarders would follow COVID-19 appropriate behaviour. Social distancing is impractical in hostels. Therefore, hostel-boarders are at an increased perceived risk of COVID-19 infection and hence their increased anxiety, corroborating with a study conducted in China.^25^

The current study also found that participants travelling to their colleges by own car had significantly greater scores for anxiety than those availing public transport. Those travelling by own cars were probably following avoidant coping mechanism by avoiding the crowd of public transport. However, those availing public transport were more accepting of the ‘new normal’ and were likely following acceptant coping mechanism. This is consistent with a study which found that psychological burden was higher with students having avoidant coping styles.^14,26^

GAD-7 scores were significantly higher in participants who had themselves suffered or had experienced their family members or relatives suffering or dying from COVID-19.^15,21^

Substance use and presence of comorbidities were not significantly associated with psychological distress, contradicted by earlier studies.^14,27^ Present study found no significant difference in symptoms of depression and anxiety among males and females, similar to a study in Bangladesh.^20^ However, some studies have found females to be more prone to psychological distress.^3^ These differences might be due to difference in characteristics of the study population based on country and culture.

The mean FCV-19S score for both study groups was lower as compared to a study conducted in India one month post lockdown.^15^ Fear of COVID-19 was neither significantly impacted by any of the socio-demographic correlates nor by academic course, compliant with other studies.^26^ This is most likely because of the time elapsed since start of the pandemic. The decrease in number of hospitalizations, deaths due to COVID-19 and pan-India vaccination might have helped in allaying fear of getting infected. However, raised levels of depression and anxiety can be attributed to the indirect effects of COVID-19. The pandemic had led to several drastic changes. Students were subjected to uncertainties regarding academics and even, their household economic condition.^24^ With gradual resumption of offline mode of teaching, the shift from their quiet life at home to hectic life at campus was hard to bear for many.^28^ This might have led students to face other sources of mental distress, or amplified their existing problems.

### Limitations and Recommendations

Since this study employed cross-sectional study design, trends of mental distress in the student population due to COVID-19 could not be assessed. Cause and effect relationships could not be established. Being a web-based, self-reported survey there could have been response bias. Non-probability sampling methods like, purposive and snowball sampling were used in this study to locate a specific student sample for comparison. However, these techniques are subjective and might have led to community bias and limited generalizability. Although standardized questionnaires were used, full diagnostic interviews were not conducted. PHQ-9 and GAD-7 questionnaires belonging to the pre-COVID era were used to assess distress during COVID-19. Information about COVID-19 vaccination was not collected. As the study was conducted among university students, its results cannot be extrapolated to the general population and healthcare professionals. While the questions asked about COVID-19 worries touched on clinical, academic and health concerns, they were not exhaustive. Thus, broader impact of the pandemic on the minds of students might have been missed. Future studies should employ better analytical study design and aim for better generalizability of sample with more detailed questionnaire including other stressors and relieving factors of the pandemic.

## CONCLUSION

Although direct harm caused by COVID-19 on mental health has been shown to improve over time,^21^ the present study has found higher prevalence of depression and anxiety than that before the pandemic in students especially, among medical students probably due to indirect and long-term consequences of the pandemic. This calls for adequate awareness and intervention. Student wellness activities like, regular sleep, balanced diet, time management, yoga, recreational activities; should be advocated in colleges across India.^29^ Positive family support is also beneficial.^30^ Students should be encouraged by college faculty to adapt to coping mechanisms like, planning, acceptance, humor, active coping, use of instrumental and emotional social support instead of gambling, substance use, avoidant coping and behavioural disengagement; which was found helpful in maintaining well-being.^26^ As students benefit most by discussing their distress with their colleagues and teammates, colleges can conduct interpersonal support groups. Student mentoring programs by faculty has also shown reduced relative prevalence of depression and anxiety in a study conducted among Indian medical students.^14^ Policy interventions such as, anti-ragging, regular mental health checkups and student grievance cells, can also help address the problem. Around the world and especially in India, seeking help from mental health professionals is met with a lot of stigma.^18^ Provision of confidential and affordable access to psychiatrists and psychologists either online or on campus, may help in mitigating this problem.

## Data Availability

All data produced in this present study are available upon reasonable request to the authors.

## ANNEXURES

### Study Questionnaire

Consent: The nature and purpose of the study and its potential risks/benefits and other relevant details of the study have been suitably explained to me in detail by the investigators. All the personal information provided by me will be kept confidential. Anonymity will be maintained. My name and email-id will not be collected. After analysis, if the result of the study be published in any article, under any circumstances, my name and identity will not be disclosed. My digital consent form indicates that I agree to participate in the study.

⎕ Yes, I agree.

#### A. Socio-demographic Characteristics

1. What is your age? ( ) years
2. What gender do you identify as?

- Male
- Female
- Other
3. What is your family size (total family members)?
4. Which of these describes your current residence?

- Home
- Hostel
- PG (paying guest)
5. Mention your course

- MBBS
- Other courses (mention:________)
6. What is the name of your college? ________________
7. Mention your year of study ____________
8. How do you travel to your college?

- Public transport
- Hired car
- Own car
- Not applicable (hostel-boarder)
9. Mention what addiction do you have?

- Tobacco
- Alcohol
- Other (specify………)
10. Do you have any prior history of psychiatric illness?

- No
- Yes
11. Please specify the disease, if you answered ‘Yes’ regarding prior history of psychiatric illness. If not, type ‘No’. __________________
12. Have you/any family member/close relative or friend, suffered from COVID-19/died of it?

- Yes (suffered/died)
- No
13. Do you have any family member who is working in health care system?

- Yes
- No
14. What morbidity do you have? (May choose more than one)

- Cardiovascular
- Respiratory e.g. Asthma
- Kidney/liver diseases
- Endocrinal/metabolic e.g. Diabetes mellitus/ thyroid dysfunction
- Cancers for which you are taking treatment/medicines
- Others (specify… )
- Nil

#### B. Specific Information

##### 1. Baseline [Knowledge, Attitude and Practice relating to COVID-19]

|  | Knowledge items | Yes | No | Do not know |
| --- | --- | --- | --- | --- |
| Sl No. | **Are the following facts correct?** |  |  |  |
| K1 | The main clinical symptoms of COVID-19 are fever, fatigue, dry cough, and myalgia (muscle pain). | 1 | 2 | 3 |
| K2 | There currently is no effective cure for COVID-2019, but early symptomatic and supportive treatment can help most patients recover from infection. | 1 | 2 | 3 |
| K3 | Not all persons with COVID-2019 will develop severe cases. Only those who are elderly have chronic illnesses are more likely to be in severe cases. | 1 | 2 | 3 |
| K4 | Eating or contacting wild animals would result in infection by the COVID-19 virus. | 1 | 2 | 3 |
| K5 | The COVID-19 virus spreads via respiratory droplets of infected individuals. | 1 | 2 | 3 |
| K6 | Ordinary residents can wear general medical masks to prevent infection by the COVID-19 virus. | 1 | 2 | 3 |

[Attitudes]
|  | **Perceived Risk of COVID-19 infection Items** | **Very low** |  | **Neither low nor high** |  | **Very high** |
| --- | --- | --- | --- | --- | --- | --- |
| A 1 | What do you think is the possibility of your COVID-19 infection? | 1 | 2 | 3 | 4 | 5 |
| A 2 | What do you think will be the severity if COVID-19 infects you? | 1 | 2 | 3 | 4 | 5 |

|  | **Efficacy beliefs Items** | **Not at all** |  |  | **Extremely** |
| --- | --- | --- | --- | --- | --- |
| **To what extent do you think the precautionary behaviour is an effective way to reduce the risk of COVID-19 infection?** |  |  |  |  |  |
| A 3 | Practicing personal hygiene such as wearing facial masks and hand hygiene' | 1 | 2 | 3 | 4 |
| A 4 | Social distancing such as avoiding crowded places. | 1 | 2 | 3 | 4 |

[Practices]
|  | **Practices of preventive behaviour** | **Never** | **Sometimes** | **Often** | **Always** |
| --- | --- | --- | --- | --- | --- |
| **In the last week, how often did you practice the following?** |  |  |  |  |  |
| P 1 | Wearing face shield/facial masks | 1 | 2 | 3 | 4 |
| P 2 | Washing hands frequently and or using hand sanitizer | 1 | 2 | 3 | 4 |
| P 3 | Avoiding visit to crowded places | 1 | 2 | 3 | 4 |

##### 2. FEAR OF COVID-19 SCALE, 2020

Please respond to each item by ticking (*√*) one of the five (5) responses that reflects how you feel, think or act toward COVID-19.

|  |  |  |  |  |  |
| --- | --- | --- | --- | --- | --- |
| Fear of COVID-19 Items | Strongly | Disagree | Neutra | Agree | Strongl |

|  |  | Disagree | e | l |  | y Agree |
| --- | --- | --- | --- | --- | --- | --- |
| 1 | I am most afraid of Corona |  |  |  |  |  |
| 2 | It makes me uncomfortable to think about Corona |  |  |  |  |  |
| 3 | My hands become clammy when I think about Corona |  |  |  |  |  |
| 4 | I am afraid of losing my life because of Corona |  |  |  |  |  |
| 5 | When I watch news and stories about Corona on Social media, I become nervous or anxious. |  |  |  |  |  |
| 6 | I cannot sleep because I'm worrying about getting Corona. |  |  |  |  |  |
| 7 | My heart races or palpitates when I think about getting Corona. |  |  |  |  |  |

#### 3. PATIENT HEALTH QUESTIONNAIRE (PHQ-9)

Over the last 2 weeks, how often have you been bothered by any of the following problems? (Use ‘*√’*eto indicate your answer)

| S.no |  | Not at all | Several days | More than half the days | Nearly every day |
| --- | --- | --- | --- | --- | --- |
| 1 | Little interest or pleasure in doing things | 0 | 1 | 2 | 3 |
| 2 | Feeling down, depressed, or hopeless | 0 | 1 | 2 | 3 |
| 3 | Trouble falling or staying asleep, or sleeping too much | 0 | 1 | 2 | 3 |
| 4 | Feeling tired or having little energy | 0 | 1 | 2 | 3 |
| 5 | Poor appetite or overeating | 0 | 1 | 2 | 3 |
| 6 | Feeling bad about yourself- or that you are a failure or have let yourself or your family down | 0 | 1 | 2 | 3 |
| 7 | Trouble concentrating on things, such as reading the newspaper or watching television | 0 | 1 | 2 | 3 |
| 8 | Moving or speaking so slowly that other people could have noticed. Or the opposite- being so fidgety or restless that you have been moving a lot more than usual | 0 | 1 | 2 | 3 |
| 9 | Thoughts that you would be better off dead, or hurting yourself | 0 | 1 | 2 | 3 |

10. If you have checked off any problems, how difficult have these problems made it for you to do your work, take care of things at home, or get along with other people?

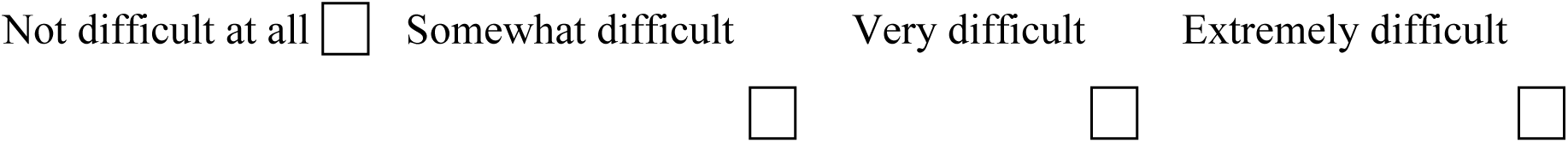

##### 4. GENERALIZED ANXIETY DISORDER SCALE (GAD-7)

| Over the <u>last two weeks</u>, how often have you been bothered by the following problems? | Not at all | Several days | More than half the days | Nearly every day |
| --- | --- | --- | --- | --- |
| 1. Feeling nervous, anxious, or on edge | 0 | 1 | 2 | 3 |
| 2. Not being able to stop or control worrying | 0 | 1 | 2 | 3 |
| 3. Worrying too much about different things | 0 | 1 | 2 | 3 |
| 4. Trouble relaxing | 0 | 1 | 2 | 3 |
| 5. Being so restless that it is hard to sit still | 0 | 1 | 2 | 3 |
| 6. Becoming easily annoyed or irritable | 0 | 1 | 2 | 3 |
| 7. Feeling afraid, as if something awful might happen | 0 | 1 | 2 | 3 |

If you checked any problems, how difficult have they made it for you to do your work, take care of things at home, or get along with other people?

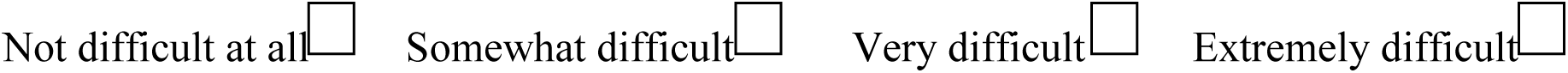

